# Validation of a Mitochondrial Polygenic Score for Parkinson’s Disease

**DOI:** 10.1101/2025.03.18.25323898

**Authors:** Joshua Chin Ern Ooi, Yi Wen Tay, Ai Huey Tan, Chin-Hsien Lin, Kajsa Atterling Brolin, Björn-Hergen Laabs, Sebastian Sendel, Inke R König, Amke Caliebe, Carolin Gabbert, Katherine M Andersh, Lietsel Jones, Lara Mariah Lange, Brian Fiske, Carolyn Sue, Christine Klein, Joanne Trinh, Theresa Lüth, Global Parkinson’s Genetics Program (GP2)

## Abstract

**Background:** Mitochondrial dysfunction is a key player in Parkinson’s disease (PD) pathogenesis. Mitochondrial polygenic scores (MGS) may be associated with PD but require validation across diverse populations.

**Objective:** To validate the association between the MGS, PD status and age-at-onset (AAO) in idiopathic and *LRRK2-*PD across various ancestries.

**Methods:** We analyzed data from 17,129 PD patients and 13,872 healthy individuals across 10 ancestries within the Global Parkinson’s Disease Genetic Program. We used regression models to assess the association between MGS, PD status and AAO.

**Results:** The MGS was associated with iPD in Europeans (β=0.19, SE=0.02, p<2.0×10^−16^) and Ashkenazi Jews (β=0.26, p=3.7×10-4) but not in other populations. Additionally, the MGS was strongly associated with *LRRK2*-PD status (β=0.82, p=2.0×10^−16^). No associations with AAO were observed.

**Conclusions:** The MGS is robustly associated with iPD status in Europeans and Ashkenazi Jews and with LRRK2-PD status. Population-specific MGS are needed to improve accuracy in other ancestries.

## Introduction

Parkinson’s disease (PD) is a complex neurodegenerative disorder with diverse symptoms.(1) As the fastest-growing neurological disease, it urgently requires effective disease-modifying therapies.(2,3) Therefore, advancing our understanding of PD pathophysiology and its phenotypic variability is essential.(1,4,5)

Mitochondrial dysfunction has long been implicated in certain forms of familial and sporadic PD.(6–10) The discovery of monogenic PD forms caused by variants in genes directly associated with mitochondrial processes (e.g. *PRKN, PINK1*, PARK7) reinforces this connection.(11–14) Furthermore, *LRRK2* mutations, the most common known causes of monogenic PD, are also linked to mitochondrial dysfunction through pathways involving mitochondrial dynamics, bioenergetics, and oxidative stress.(15)

Genome-wide association studies (GWAS) have improved insights into the genetic architectures of complex diseases.(10,16,17) Polygenic scores leverage these data by capturing the cumulative impact of multiple variants implicated in a disease. They provide a framework to explore genetic contributions to pathomechanisms underlying sporadic and monogenic PD.(16,18–22) However, most neurodegenerative disorder GWASs (82%) are based on European cohorts, thereby limiting our knowledge of PD genetics in diverse populations.(23,24)

Efforts have focused on developing mitochondrial polygenic scores (MGS), which quantify the cumulative impact of mitochondrial function-associated variants.(10) Although some MGS show associations with PD status,(25,26) replication in large, independent cohorts remains limited.(27) Moreover, population-specific differences in genetic and environmental factors may influence MGS efficacy across ancestries.(26)

We recently developed an MGS using European individuals, demonstrating its association with idiopathic PD (iPD) status and age-at-onset (AAO) in *LRRK2*-PD.(26) Here, we aim to replicate and expand our findings by evaluating its association with PD status and AAO for both iPD and *LRRK2*-PD across a large, multi-ancestry cohort.

## Methods

### Study Dataset and Populations

We used genetic and clinical data from the Global Parkinson’s Disease Genetic Program (GP2),(28) release 6 dataset (https://doi.org/10.5281/zenodo.10472143). This includes imputed genotype data generated using the NeuroBooster array,(29) with quality control conducted using GenoTools,(30) to prune call rates and assess discordant sex, duplicates, relatedness, and heterozygosity (https://github.com/GP2code/GenoTools).(30)

Ancestry was estimated using a diverse reference panel, and imputed through *TOPMed* (https://topmed.nhlbi.nih.gov/).(31,32) Principal component analysis and clustering using ancestry-specific genetic markers categorized a total of 17,129 PD patients and 13,872 healthy individuals into 11 genetically determined ancestries—African, African admixed, Ashkenazi Jewish, Latino and Indigenous people of the Americas, East Asian, European, South Asian, Central Asian, Middle Eastern, Finnish, and Complex Admixture (**Table 1**).

**Table 1:** Association between Mitochondrial Polygenic Scores and Parkinson’s Disease Status.

| Ancestry | Patients | Controls | $\beta$ | Standard Error | p-value |
| --- | --- | --- | --- | --- | --- |
| **Idiopathic PD** |  |  |  |  |  |
| EUR | 11,739 | 7,596 | 0.19 | 0.02 | $< 2 \times 10^{-16} *$ |
| AJ | 864 | 459 | 0.26 | 0.07 | $3.7 \times 10^{-4} *$ |
| MDE | 202 | 21 | 0.48 | 0.27 | 0.077 |
| AFR | 918 | 1,677 | 0.07 | 0.09 | 0.402 |
| AMR | 354 | 139 | 0.18 | 0.12 | 0.130 |
| AAC | 256 | 806 | 0.05 | 0.09 | 0.594 |
| EAS | 1,576 | 2,378 | 0.07 | 0.05 | 0.138 |
| CAS | 285 | 294 | 0.13 | 0.11 | 0.238 |
| CAH | 440 | 280 | 0.09 | 0.10 | 0.338 |
| SAS | 122 | 207 | -0.10 | 0.19 | 0.597 |
| **LRRK2-PD\*** |  |  |  |  |  |
| combined | 282 | 13,844 | 0.82 | 0.06 | $< 2 \times 10^{-16} *$ |

**Legend:** Table detailing the number of patients and healthy individuals and association between MGS and PD status. EUR: European, AJ: Ashkenazi Jewish, MDE: Middle Eastern, AFR: African, AMR: Latino and Indigenous People of the Americas, AAC: African Admixed, EAS: East Asian, CAS: Central Asian, CAH: Complex Admixture, SAS: South Asian, Patients: Number of patients with idiopathic PD (excluding patients with *PINK1/PRKN/LRRK2*-PD) or *LRRK2*-PD, $\beta$ : Regression coefficient, \*: $p < 0.005$ (significance levels were adjusted according to the Bonferroni method to $\alpha = 0.005$ for multiple testing corrections ( $n=10$ populations)), \*: All ancestry groups pooled together, Regression model: $\text{logit}(P(\text{CASE}=1)) = \beta_0 + \beta_1(\text{standardized MGS score}) + \beta_2(\text{sex}) + \beta_3(\text{PC1}) + \dots + \beta_{10}(\text{PC10}) + \beta_{11}(\text{age})$

Further information on GP2 data acquisition, imputation, quality control, and release policies are available at https://gp2.org/.

Of the patients, 62.3% were male. AAO data was available for 10,348 patients, with a mean AAO of 57.0 years (SD=12.6 years), and disease duration (Age-at-baseline - AAO) of 9.0 years (SD=7.1 years). **Supplementary Table 1** summarizes demographics by ancestry. In this study, populations with fewer than 100 individuals (i.e. Finnish) were excluded.

### Mitochondrial Polygenic Score Generation and Analysis

We previously developed an MGS using the “ProtectMove” cohort (http://protect-move.de/), comprising European individuals distinct from the GP2 database.(26) Leveraging summary statistics from the Nalls *et al. G*WAS,(16) and focusing on SNPs associated with mitochondrial function outlined by Billingsley *et al*.,(10) we applied five-fold cross-validation using *PRSice-2*,(33) *LDpred2*,(34) and *lassosum2*.(35)

Our resultant MGS comprised 14,789 SNPS and the effect alleles with corresponding weights are available at https://github.com/LuethTheresa/MitochondrialPolygenicScoreAndAgeAtOnset. It achieved an area under the receiver operating curve (AUC) of 0.56 (95%CI=0.54—0.58) and an odds ratio of 1.25 per standard deviation (SD). Further details on the development of this MGS are in **Supplementary Text 1** and our previous publication.(26)

In this study, using GP2 data, we applied additional quality control measures, including minor allele frequency > 0.01, missingness per sample < 0.02, missingness per SNP < 0.05, and Hardy-Weinberg equilibrium p-value > 1×10^−50^. Individual MGS were calculated using PLINK (v1.9/v2.0) and standardized (mean=0, SD=1).(36) MGS SNP percentages by ancestry are detailed in **Supplementary Table 2**.

**Table 2:** Association between Mitochondrial Polygenic Scores and Age-at-Onset in Patients with Idiopathic PD.

| Ancestry | N | $\beta$ | Standard Error | p-value |
| --- | --- | --- | --- | --- |
| **Idiopathic PD** |  |  |  |  |
| EUR | 7,492 | -0.26 | 0.14 | 0.058 |
| AJ | 690 | -0.19 | 0.43 | 0.663 |
| MDE | 145 | 0.18 | 0.94 | 0.851 |
| AFR | 159 | -0.54 | 1.09 | 0.619 |
| AMR | 246 | -0.57 | 0.86 | 0.511 |
| AAC | 149 | 0.30 | 1.05 | 0.774 |
| EAS | 726 | -0.29 | 0.43 | 0.502 |
| CAS | 114 | 2.09 | 1.35 | 0.126 |
| CAH | 296 | -0.62 | 0.84 | 0.457 |
| SAS | 66 | 1.67 | 2.09 | 0.429 |
| ***LRRK2*-PD\*** |  |  |  |  |
| combined | 229 | 0.29 | 0.85 | 0.735 |

**Legend:** Table detailing the number of patients and healthy individuals and association between MGS and age-at-onset for idiopathic PD and *LRRK2*-PD. EUR: European, AJ: Ashkenazi Jewish, MDE: Middle Eastern, AFR: African, AMR: Latino and Indigenous People of the Americas, AAC: African Admixed, EAS: East Asian, CAS: Central Asian, CAH: Complex Admixture, SAS: South Asian, N: Number of idiopathic PD patients (excluding patients with *PRKN*/*PINK1*/*LRRK2*-PD) or *LRRK2*-PD patients with age-at-onset data available, $\beta$ : regression coefficient, \*: All ancestry groups pooled together, Regression model: $AAO = \beta_0 + \beta_1(\text{standardized MGS score}) + \beta_2(\text{sex}) + \beta_3(PC1) + \dots + \beta_{10}(PC10)$

### Genetic Stratification of PD Subgroups

*P*atients with pathogenic *LRRK2 v*ariants (**Supplementary Table 3**), as determined by Krüger *et al*.,(37) were identified and classified as *LRRK2-*PD. Individuals homozygous or compound heterozygous for pathogenic *PRKN/PINK1 v*ariants (**Supplementary Table 4**) were removed and the remaining patients were classified as iPD. The low number of *PRKN/PINK1-*PD patients precluded further analyses in this group.

### Statistical Analyses

*A*nalyses and visualizations were performed using *R (*v4.4.0).(38) Normality was confirmed before performing t-tests. The association between MGS and disease status was assessed using multivariable logistic regression, adjusting for sex, age-at-baseline, and principal components (PCs 1-10). Power calculations performed based on our previous study’s effect size, indicated that 484 iPD patients and 238 healthy individuals were required to achieve 80% power.(26) AUC analyses assessed predictive accuracy. For patients, the relationship between MGS and AAO was examined using linear regression models, adjusting for sex and PCs 1-10. All iPD-related analyses were stratified by ancestry groups.

Given our *a priori h*ypothesis of an association between MGS and PD status, significance was set at α = 0.005 for the ten evaluated ancestries (Bonferroni correction: 0.05/10). All other analyses on MGS and AAO remained exploratory, with uncorrected p-values. All tests were two-sided.

Analyses were performed on the Terra Community Workbench (https://app.terra.bio/).

## Results

### Association between MGS and iPD status

*I*n the European group, iPD patients had a higher MGS (MGS=0.18, SD=1.00) than healthy individuals (MGS=0.00, SD=1.00, t-test: p<2.2×10^−16^, **Figure 1A**). Similarly, in the Ashkenazi Jewish group, iPD patients had higher MGS (MGS=0.21, SD=1.02) compared to healthy individuals (MGS=0.00, SD=1.00, t-test: p=1.90 × 10^−4^, **Figure 1B**). Multivariable logistic regression confirmed the significant association between higher MGS and iPD status in both groups (European: β=0.19, SE=0.02, p<2.0×10^−16^, Ashkenazi Jewish: β=0.26, SE=0.07, p=3.7×10^− 4^)(**Table 1**). No associations were observed in other ancestry groups. The AUC was 0.55 (95%CI=0.54—0.56) and 0.56 (95%CI=0.53—0.59) in the Europeans and Ashkenazi Jewish, respectively (**Figure 1D-E**).

**Figure 1:**
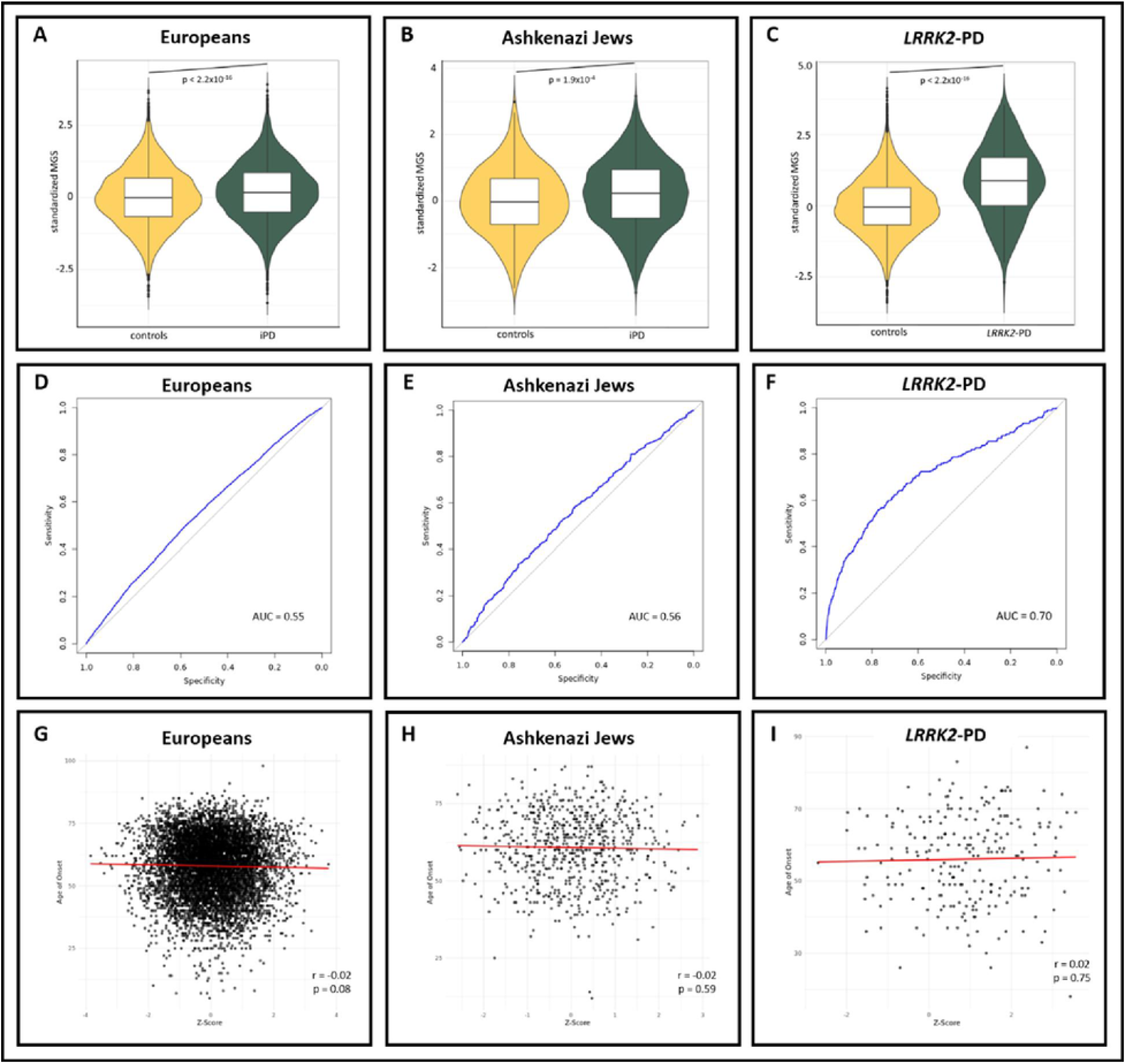
Associations between Mitochondrial Polygenic Scores and Parkinson’s Disease Status and Age-at-Onset. **(A-C):** Violin plots show the distribution of standardized MGS for patients with idiopathic PD (iPD) and healthy individuals in the European **(A)** and Ashkenazi Jewish **(B)** ancestry groups, as well as for patients with *LRRK2-PD* ***(C)***. *Bo*xplots within the violins indicate the median (solid line) and interquartile range (box edges), while the violin shape reflects the overall data distribution. p = t-test p-value. **(D-E**): Receiver operating curves and respective area under curve values for European **(D)** and Ashkenazi Jewish **(E)** ancestry groups, as well as patients with *LRRK2-PD* ***(F). (G*-I)**: Correlation plots display the association between MGS and AAO in patients with iPD in the European **(G)** and Ashkenazi Jewish **(H)** ancestry groups, as well as patients with *LRRK2-PD* ***(I)***. *r* = Spearman’s rank correlation coefficient, p = Spearman’s exploratory p-value.

### Association between MGS and AAO in iPD

*I*n the European group, we observed a weak inverse trend between a higher MGS and an earlier AAO for iPD (r=-0.02, p=0.08, **Figure 1G**). A similar result was also seen in the Ashkenazi Jews (r=-0.02, p=0.59, **Figure 1H**). Multivariable regression revealed a trend for an inverse association between MGS and AAO in the European group (β=-0.26, SE=0.14, p=0.058), with no associations detected in the other populations (**Table 2**).

### Analyses on LRRK2-PD

*G*iven the limited number of *LRRK2 v*ariant carriers within each ancestry group (**Supplementary Table 3**), we pooled all *LRRK2-*PD patients (N=282) and non-*LRRK2 v*ariant-carrying healthy individuals (N=13,844) across ancestries for a combined analysis. We observed a higher MGS in *LRRK2-*PD patients (MGS=0.84, SD=1.24) compared to controls (MGS=0.00, SD=1.00, t-test: p<2.2×10^−16^, **Figure 1C**) and an AUC of 0.70 (95%CI=0.67—0.74)(**Figure 1F**). Multivariable logistic regression showed a strong association between MGS and *LRRK2-*PD status (ß=0.82, SE=0.06, p=2.07×10^−16^, **Table 1**). No association was observed between MGS and AAO in *LRRK2-* PD (ß=0.29, SE=0.85, p=0.735, **Table 2**).

To address the disproportionate ratio of *LRRK2-*PD patients to controls, we weighted patients to healthy individuals at 1:3 and down-sampled the AUC analysis, both yielding similar results. A sub-analysis focusing solely on European and Ashkenazi Jewish individuals, which comprised the majority of *LRRK2-*PD patients, similarly showed consistent associations. Results are in **Supplementary Figures 1&2**.

## Discussion

We previously developed an MGS that was associated with iPD status in the Accelerating Medicines Partnership®-Parkinson’s disease cohort.(26) In this study, using GP2 data, we replicated this finding in European and Ashkenazi Jewish populations, confirming its validity. Its performance in distinguishing iPD patients from healthy individuals (European: AUC=0.55, Ashkenazi Jewish: AUC=0.56) was consistent with our previous findings (AUC=0.56).(26)

The lack of significant MGS associations in African and East Asian populations, despite adequate cohort sizes, likely reflects differences in genetic architecture. Derived from European data, the MGS may be biased toward variants more prevalent in Europeans and related populations. Only 78.2% and 58.9% of the 14,789 MGS SNPs are represented in African and East Asian populations, respectively, compared to 88.0% in Europeans and 95.9% in Ashkenazi Jews. These disparities highlight inherent differences in allele frequencies, variant distributions, and cross-population overlaps.

Differences in linkage disequilibrium (LD) structure may also contribute. As most GWAS variants tag nearby causal variants rather than being causal themselves, varying LD patterns across ancestries may reduce SNPs effectiveness in non-European populations. Lower SNP representation might also stem from monomorphism, absence from genotyping arrays, or QC filtering. These emphasize the need for population-specific MGS.

Nonetheless, the under-representation of non-European ancestries in GP2’s dataset remains a limitation. Most other ancestry groups fell short of the 484 iPD patients and 238 healthy individuals required to achieve 80% power (**Table 1**). Expanding these under-represented groups could uncover associations missed here, highlighting the need for inclusive genomic studies.(24)

We observed a weak trend for an inverse association between MGS and AAO in Europeans. The stronger association between MGS and iPD status compared to AAO might suggest that different molecular pathways govern these processes.

The strong association between MGS and *LRRK2-*PD status in our pooled analysis reinforces the link between *LRRK2-*PD and mitochondrial dysfunction.(24) The MGS showed stronger predictive utility for *LRRK2-*PD (AUC=0.75) than iPD (AUC=0.55-0.56), suggesting it more effectively captures mitochondrial dysfunction in this monogenic form, whereas iPD status could be influenced by greater genetic and environmental heterogeneity.

No association was found between MGS and AAO in *LRRK2-*PD, likely due to sample size limitations and heterogeneity within *LRRK2 c*arriers. Approximately one-third of *LRRK2-*PD patients lack neuronal-predominant misfolded and aggregated alpha-synuclein, suggesting that *LRRK2-*PD represents a spectrum.(39) Stronger MGS associations may emerge within specific subgroups, such as alpha-synuclein-negative *LRRK2-*PD, reflecting distinct underlying mechanisms. Larger, stratified studies may clarify these dynamics and further elucidate mitochondrial dysfunction’s role in modulating disease onset and progression across PD subtypes, including other monogenic forms like *PRKN/PINK1-*PD, which were under-represented in this dataset.

Examining how environmental exposures and lifestyle factors interact with MGS,(26) and how these gene-environment interactions vary across populations will be valuable. While current limitations in environmental data within GP2 restrict such analyses, comprehensive efforts are underway to address these gaps.

In conclusion, we validated the association between our MGS and iPD status in European and Ashkenazi Jewish populations and observed a strong association with *LRRK2-*PD status. Our results also highlight the need for population-specific MGS and for more comprehensive phenotyping and environmental data to fully understand the interactions between genetic and non-genetic factors in PD globally. Continued efforts in these areas will enhance our understanding of PD, ultimately leading to more effective treatment strategies.

## Acknowledgements

Data used in the preparation of this article were obtained from Global Parkinson’s Genetics Program (GP2). GP2 is funded by the Aligning Science Across Parkinson’s (ASAP) initiative and implemented by The Michael J. Fox Foundation for Parkinson’s Research (https://gp2.org). For a complete list of GP2 members see https://gp2.org.

## Data Availability Statement

Data sharing is not applicable to this article as no new data were created or analyzed in this study. Data used in the preparation of this manuscript were obtained from the Global Parkinson’s Genetics Program (GP2) database accessed via the Terra platform (https://app.terra.bio/#workspaces). For up-to-date information on GP2 data acquisition, access, and policies, visit https://gp2.org/.

All code generated for this article, and the identifiers for all software programs and packages used, are available on GitHub (https://github.com/GP2code/GP2-mitochondrial-PRS) and were given a persistent identifier via Zenodo (https://doi.org/10.5281/zenodo.15013765).

## Funding acknowledgement

This project was supported by the Global Parkinson’s Genetics Program (GP2; https://gp2.org). GP2 is funded by the Aligning Science Across Parkinson’s (ASAP) initiative and implemented by The Michael J. Fox Foundation for Parkinson’s Research (MJFF). For a complete list of GP2 members see doi.org/10.5281/zenodo.7904831

## Conflict(s) of Interest

| **Author** | **COI** |
| --- | --- |
| **JCEO** | Nothing to declare |
| **YWT** | Nothing to declare |
| **AHT** | Nothing to declare |
| **CHL** | Nothing to declare |
| **KAB** | Nothing to declare |
| **BHL** | Nothing to declare |
| **SS** | Nothing to declare |
| **IRK** | Nothing to declare |
| **AC** | Nothing to declare |
| **CG** | Nothing to declare |
| **KA** | Nothing to declare |
| **LJ** | Nothing to declare |
| **LML** | Nothing to declare |
| **BF** | Nothing to declare |
| **CS** | Nothing to declare |
| **CK** | Nothing to declare |
| **TL** | Nothing to declare |
| **JT** | Nothing to declare |

