## Supplementary for "Validation of a Mitochondrial Polygenic Score for Parkinson’s Disease"

**Supplementary Material**

**Supplementary Table 1: Demographics of Included GP2 Participants**

|  | | **Controls** | **iPD** |
| --- | --- | --- | --- |
| **All Ancestries Combined** | N | 13,872 | 17,129 |
|  | Male (%) | 58.0 | 62.3 |
|  | Mean AAB [SD] (years) | 63.7 [12.2] | 66.2 [11.2] |
|  | Mean AAO [SD] (years) |  | 57.0 [12.6] |
| **AAC** | N | 808 | 257 |
|  | Male (%) | 36.6% | 55.3% |
|  | Mean AAB [SD] (years) | 65.9 [10.2] | 65.7 [11.3] |
|  | Mean AAO [SD] (years) |  | 58.5 [12.5] |
| **AFR** | N | 1,683 | 920 |
|  | Male (%) | 50.0 | 66.8 |
|  | Mean AAB [SD] (years) | 63.6 [15.3] | 63.0 [12.1] |
|  | Mean AAO [SD] (years) |  | 57.1 [12.8] |
| **AJ** | N | 459 | 1,011 |
|  | Male (%) | 64.7 | 70.2 |
|  | Mean AAB [SD] (years) | 67.7 [9.7] | 69.3 [9.9] |
|  | Mean AAO [SD] (years) |  | 60.2 [11.6] |
| **AMR** | N | 139 | 367 |
|  | Male (%) | 48.9 | 57.2 |
|  | Mean AAB [SD] (years) | 63.2 [9.0] | 59.7 [12.0] |
|  | Mean AAO [SD] (years) |  | 50.2 [13.9] |
| **CAH** | N | 281 | 452 |
|  | Male (%) | 45.9 | 55.8 |
|  | Mean AAB [SD] (years) | 46.3 [20.1] | 61.2 [12.6] |
|  | Mean AAO [SD] (years) |  | 53.5 [14.5] |
| **CAS** | N | 294 | 286 |
|  | Male (%) | 51.0 | 50.7 |
|  | Mean AAE [SD] (years) | 54.4 [6.62] | 58.4 [12.2] |
|  | Mean AAO [SD] (years) |  | 46.7 [13.7] |
| **EAS** | N | 2,378 | 1,577 |
|  | Male (%) | 72.5 | 55.0 |
|  | Mean AAB [SD] (years) | 62.4 [11.2] | 66.1 [10.6] |
|  | Mean AAO [SD] (years) |  | 50.3 [13.3] |
| **EUR** | N | 7,596 | 11,841 |
|  | Male (%) | 57.8 | 63.2 |
|  | Mean AAB [SD] (years) | 64.9 [10.7] | 66.6 [10.9] |
|  | Mean AAO [SD] (years) |  | 57.9 [12.1] |
| **FIN** | N | 6 | 78 |
|  | Male (%) | 33.3 | 57.7 |
|  | Mean AAB [SD] (years) | 80.0 [7.5] | 64.7 [10.7] |
|  | Mean AAO [SD] (years) |  | 55.2 [11.3] |
| **MDE** | N | 21 | 218 |
|  | Male (%) | 76.2 | 64.7 |
|  | Mean AAB [SD] (years) | 67.5 [13.6] | 64.5 [12.8] |
|  | Mean AAO [SD] (years) |  | 55.1 [13.3] |
| **SAS** | N | 207 | 122 |
|  | Male (%) | 68.1 | 68.0 |
|  | Mean AAB [SD] (years) | 54.6 [16.9] | 62.6 [13.5] |
|  | Mean AAO [SD] (years) |  | 55.1 [13.3] |
| ***LRRK2* Variant Carriers** | | | |
| **All Ancestries Combined** |  | Unaffected Variant Carriers | *LRRK2*-PD |
|  | N | 23 | 282 |
|  | Male (%) | 65.2 | 55.7 |
|  | Mean AAB [SD] (years) | 57.3 [8.0] | 67.3 [11.4] |
|  | Mean AAO [SD] (years) |  | 56.1 [12.2] |

**Legend:** EUR: European, AJ: Ashkenazi Jewish, MDE: Middle Eastern, AFR: African, AMR: Latino and Indigenous People of the Americas, AAC: African Admixed, EAS: East Asian, CAS: Central Asian, CAH: Complex Admixture, SAS: South Asian, N: Number of individuals, AAB: Age-at-baseline, AAO: Age-at-onset, PD: Parkinson's disease, *LRRK2*-PD: Patients with PD that carry a *LRRK2* variant

**Supplementary Table 2: Numbers and Percentages of MGS SNPs Identified in Each Ancestry Group**

| **Ancestry** | **N** | **%** |
| --- | --- | --- |
| **EUR** | 13,012 | 88.0 |
| **AJ** | 14,178 | 95.9 |
| **MDE** | 12,248 | 82.8 |
| **AFR** | 11,564 | 78.2 |
| **AMR** | 13,656 | 92.3 |
| **AAC** | 13,752 | 93.0 |
| **EAS** | 8,716 | 58.9 |
| **CAS** | 12,464 | 84.3 |
| **CAH** | 13,194 | 89.2 |
| **SAS** | 12,163 | 82.2 |

**Legend:** AAC: African Admixed, AFR: African, AJ: Ashkenazi Jewish, AMR: Latino and Indigenous People of the Americas, CAS: Central Asian, CAH: Complex Admixture, EAS: East Asian, EUR: European, MDE: Middle Eastern, SAS: South Asian, N: Number of MGS SNPs, %: Percentage of the total 14,789 SNPs that were present

**Supplementary Table 3: *LRRK2* Variant List and Corresponding Number of Patients in Each Ancestry Group**

| **Variant** | **Chr:Position (hg38)** | **N** | | | | | | | | | |
| --- | --- | --- | --- | --- | --- | --- | --- | --- | --- | --- | --- |
|  |  | **AAC** | **AFR** | **AJ** | **AMR** | **CAS** | **CAH** | **EAS** | **EUR** | **MDE** | **SAS** |
| **A1442P** | chr12:40310437:G:C | 0 | 0 | 0 | 0 | 0 | 0 | 0 | 0 | 0 | 0 |
| **G2019S** | chr12:40340400:G:A | 0 | 0 | **145** | **12** | 0 | **8** | 0 | **84** | **11** | 0 |
| **I2020T** | chr12:40340404:T:C | 0 | 0 | 0 | 0 | 0 | 0 | 0 | 0 | 0 | 0 |
| **L1795F** | chr12:40322386:G:T | 0 | 0 | 0 | 0 | 0 | 0 | 0 | 0 | 0 | 0 |
| **N1437H** | chr12:40309225:A:C | 0 | 0 | 0 | 0 | 0 | 0 | 0 | 0 | 0 | 0 |
| **R1325Q** | chr12:40308481:G:A | 0 | 0 | 0 | 0 | **1** | 0 | 0 | **7** | 0 | 0 |
| **R1441C** | chr12:40310434:C:T | 0 | 0 | 0 | 0 | 0 | 0 | 0 | **10** | **4** | 0 |
| **R1441G** | chr12:40310434:C:G | 0 | 0 | 0 | 0 | 0 | 0 | 0 | 0 | 0 | 0 |
| **R1441H** | chr12:40310435:G:A | 0 | 0 | 0 | 0 | 0 | 0 | 0 | 0 | 0 | 0 |
| **V1447M** | chr12:40310452:G:A | 0 | 0 | 0 | 0 | 0 | 0 | 0 | 0 | 0 | 0 |
| **Y1699C** | chr12:40321114:A:G | 0 | 0 | 0 | 0 | 0 | 0 | 0 | 0 | 0 | 0 |

**Legend:** AAC: African Admixed, AFR: African, AJ: Ashkenazi Jewish, AMR: Latino and Indigenous People of the Americas, CAS: Central Asian, CAH: Complex Admixture, EAS: East Asian, EUR: European, MDE: Middle Eastern, SAS: South Asian, N: number of patients with particular *LRRK2* variant, non-zero numbers are in bold

**Supplementary Table 4: List of Pathogenic/Likely Pathogenic *PRKN* and *PINK1* Variants**

| ***PRKN*** | ***PINK1*** |
| --- | --- |
| c.619-1G>A  c.1286-3C>G  p.Met1Thr  p.Gln34Argfs*5  p.Arg42Pro  p.Asn52Metfs*29  p.Asp53*  p.Val56Glu  p.Gln178*  p.Cys212Trpfs*13  p.Cys212Tyr  p.Thr240Met  p.Cys238Trp  p.Arg256Cys  p.Arg275Trp  p.Gly284Arg  p.Gln311*  p.Cys332*  p.Val324Alafs*111  p.Leu325Cysfs*110  p.Arg348Glufs*21  p.Lys349Ilefs*21  p.Gly430Asp  p.Pro437Leu  p.Cys441Arg  p.Asp460Glyfs*109 | p.Trp90*  p.Gln126Pro  p.Ala168Pro  p.Gly309Asp  p.Thr313Met  p.Gly386Ala  p.Gly409Arg  p.Leu489Pro |

**Supplementary Text 1: Generation of the Mitochondrial Polygenic Score**

This section outlines the steps involved in generating our mitochondrial polygenic score (MGS). These steps were performed in our previous study and were not repeated here.(1) We utilized samples from the Research Group “Protect Move” (FOR2488)( <http://protect-move.de/>) which is funded by the German Research Foundation DFG. After quality control, the dataset included 1,914 Parkinson’s disease (PD) patients and 4,464 controls collated from five different German cohorts. Further information of these cohorts including genotyping and quality control protocols, have been published previously.(2)

To focus on idiopathic PD, we excluded all samples with mutations in known PD-associated genes (*CHCHD2, DJ-1, LRRK2, PINK1, PRKN, SNCA, VPS35,* and *GBA1*). This resulted in a final dataset of 6,000 samples (2,800 females, 3,161 males, 39 ambiguous) comprising 1,805 patients and 4,195 healthy individuals.

For MGS development, we employed summary statistics from the GWAS performed by Nalls *et al.*,(3) with permission from 23andMe. Our analyses were restricted to SNPs located in genes associated with mitochondrial function, as defined in the “secondary gene” list by Billingsley *et al.*,(4) yielding a total of 168,629 SNPs as the basis for our MGS.

The dataset was then divided into five equally sized subsets and stratified by case-control status, sex, age at onset (patients), and age (controls). This was performed with the R package "groupdata2" (version 2.0.2) (<https://CRAN.R-project.org/package=groupdata2>).

Using these five datasets, we utilized a five-fold cross-validation approach, where the MGS was trained on four out of five of the datasets and then validated on the remaining fifth. With each of these datasets, we generated multiple MGSs using three different tools: PRSice-2,(5) LDpred2,(6) and lassosum2.(7)

- PRSice-2: All combinations resulting from window sizes 250, 500, 750, and 1000 kB and r² thresholds 0.3, 0.35, 0.4, 0.45, 0.5, 0.55, 0.6, 0.65, 0.7, 0.75, and 0.8 were utilized. Linkage disequilibrium (LD) was calculated using either the training data or the 1000 Genomes Project as a reference dataset.
- LDpred2 and lassosum2: SNP correlation matrices were calculated with the R package "bigsnpr" (version 1.9.11).(8) For LDpred2, three models ("inf", "auto", and "grid") were applied. The "auto" model varied over 30 *p* parameters logarithmically distributed between 0.0001 and 0.5. The “grid” model used a grid consisting of weights for the SNP heritability *h²* with values 0.7, 1, and 1.4 as well as 17 values for the proportion of causal variants *p* between 0.0001 and 1.0 on a logarithmic scale, for both the sparse and non-sparse versions.

After calculating the MGS models as described, the best-performing model for each training dataset and method [PRSice-2 (LD-reference own data), PRSice-2 (LD-reference 1000 genomes), LDpred2 inf, LDpred2 auto, LDpred2 grid, lassosum2] was selected based on the highest AUC. These models were then applied to their respective validation datasets, and the mean AUC was calculated across all five validation datasets per model. Among the methods, the LDpred2 auto model performed best. We therefore computed an MGS with the whole dataset as a training dataset using LDpred2 auto. This final model achieved an AUC of 0.560 (95% CI: [0.544, 0.576]) with an odds ratio of 1.245 per standard deviation.

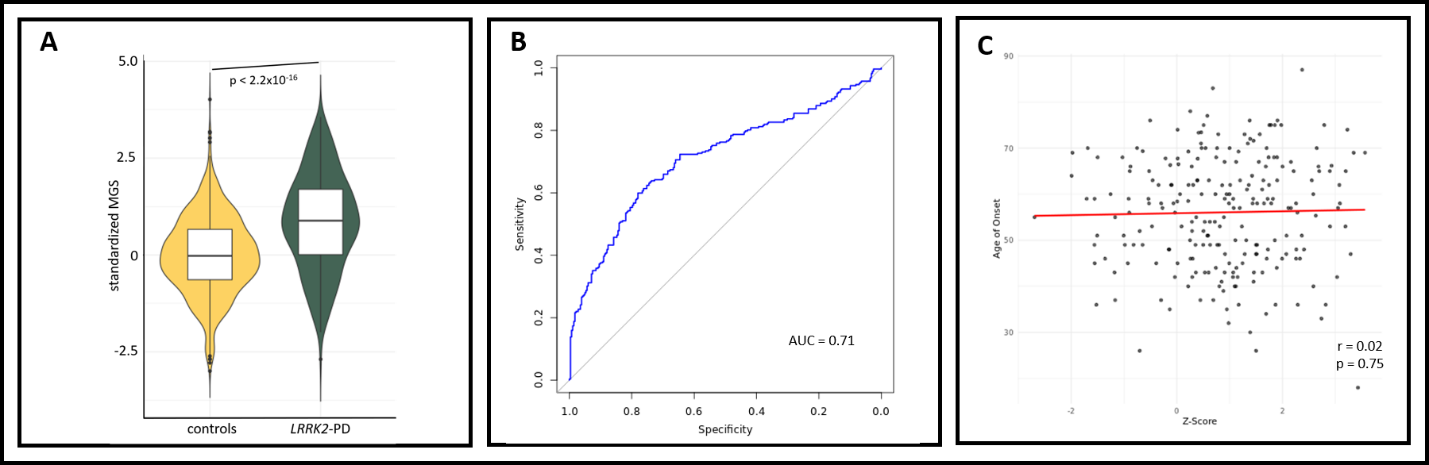

**Supplementary Figure 1: Weighted/Down-sampled *LRRK2*-PD Sub-analysis**

To account for the disproportionate ratio of *LRRK2*-PD patients to healthy individuals, we performed additional analyses weighting patients to controls at a 1:3 ratio. (A): Similarly, patients with *LRRK2*-PD had higher MGS (MGS=0.84, SD=1.24) compared to healthy individuals (MGS=0.00, SD=1.00, Welch t-test: p<2.2×10^−16^). Multivariable logistic regression showed a strong association between MGS and *LRRK2*-PD status (ß=0.80, SE=0.04, p=2.0x10^-16^). (B): Down-sampling of the AUC analysis yielded an AUC of 0.71 (95%CI=0.67—0.76). (C): Results of multivariate linear regression for MGS and AAO in *LRRK2*-PD remain unaffected.

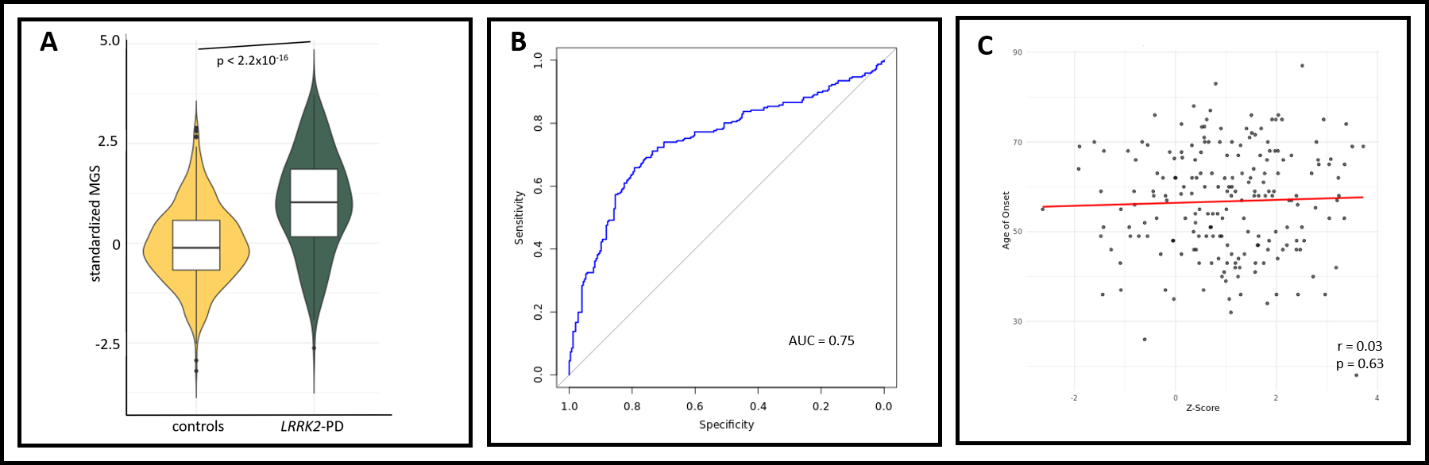
**Supplementary Figure 2: Weighted/Down-sampled *LRRK2*-PD Sub-analysis for Only Europeans and Ashkenazi Jewish Patients and Controls**

We performed further sub-analyses focusing solely on patients and healthy individuals from European and Ashkenazi Jewish ancestries, which constituted the majority of *LRRK2*-PD patients. We identified 246 *LRRK2*-PD patients and 8037 healthy individuals. Analyses were weighted and down-sampled appropriately. (A): Similarly, patients with *LRRK2*-PD had higher MGS (MGS=1.00, SD=1.26) compared to healthy individuals (MGS=0.00, SD=1.00, Welch t-test: p<2.2×10^−16^). Multivariable logistic regression showed a strong association between MGS and *LRRK2*-PD status (ß=0.85, SE=0.04, p=2.0x10^-16^). (B): Down-sampling of the AUC analysis yielded an AUC of 0.75 (95%CI=0.70—0.79). (C): Results of multivariate linear regression for MGS and AAO in *LRRK2*-PD remain unaffected.
